# A single holiday was the turning point of the COVID-19 policy of Israel

**DOI:** 10.1101/2020.03.26.20044412

**Authors:** Ziv Klausner, Eyal Fattal, Eitan Hirsch, Shmuel C. Shapira

## Abstract

**Background:** The impact of COVID-19 has been profound, and the public health challenge seem to be the most serious regarding respiratory viruses since the 1918 H1N1 influenza pandemic. In the absence of effective vaccine or biomedical treatment, the basic rules of public health measures have not changed, namely public distancing.

**Methods:** We analyzed epidemiological investigation reports during the first month of the outbreak in Israel. In addition, we present a deterministic compartment model and simulations of several scenarios emphasizing quarantine and isolation policies given their efficiency.

**Results:** We identify an abrupt change from controlled epidemic regime to an exponential growth (*R*_0_ = 2.19) in light of the actual policy-makers decisions and public behavior in Israel. Our analysis show that before the abrupt change, the new cases trend was due to returning citizens infected abroad. The abrupt change followed a holiday in which social distancing was clearly inefficient and many public gatherings were held. We further discuss three different modeled scenarios of quarantine efficiency: high-, medium-, and low-efficiency.

**Conclusions:** Israel early lessons show that there is no allowance to compromise with the directive of social distancing. Even before the onset of the pandemic in Israel, fine-tuned but determined early decisions were taken by policy makers to monitor flight arrivals from Covid-19 affected regions and to limit public gatherings. Our analysis show that one particular holiday has shifted the occurrence curve from controlled regime to exponential growth. Therefore, even a short lapse in public responsiveness can have a dramatic effect.

## 1. Introduction

Since its emergence the impact of COVID-19 has been profound, and the public health challenge seem to be the most serious seen in a respiratory virus since the 1918 H1N1 influenza pandemic (Soper 1919). In this study we present the results of epidemiological data and modelling of one month since the onset of the outbreak in Isreal, addressing public events occurring during this period and the sensitivity to a number of public health measures focusing on social distancing (quarantine and isolation).

The epidemiological data studied, consisting of 381 laboratory confirmed COVID-19 cases, has been obtained from the epidemiological investigation reports that were released by the Israeli Ministry of Health (Israeli Ministry of Health, 2020). This allowed us to identify and separate the incoming from abroad (which policy required to enter quarantine on arrival) to the local cases, thus analyzing the net contribution of local infections.

In addition, we present an extended deterministic SEIR (Susceptible, Exposed, Infectious, and Recovered) model to simulate disease outbreak scenarios. In particular, the model includes quarantine of asymptomatic suspected population (exposed) and isolation of symptomatic and infectious patients. The model takes into account the efficiency of the quarantine and isolation measures. We discuss three different quarantine efficiency scenarios: high-efficiency, medium-efficiency, and low-efficiency. The resulting analysis from the epidemiological cases data are discussed in light of public events and compared to model simulations. We analyze and discuss an abrupt change from controlled epidemic regime to an exponential growth regime in light of policy makers’ decisions and public behavior.

## 2. Methods

The dynamics of spread of epidemics as well as the quarantine-isolation policy of Israel was modeled using the SEQIJR model (e.g. Gumel, et al., 2004). This is a deterministic compartmental model which allows the implicit inclusion of biological epidemiological phases (including incubation period) as well as governmental interventions such as quarantine and their actual efficiency of implementation. A successful a posteriori implementation of this model to the transmission dynamics and control of the SARS epidemics in Toronto, Hong Kong, Singapore and Beijing is given in Gumel, et al., 2004. The model consists of a system of 7 dynamical equations and 15 parameters. For details of the model and its parameters see the online supplementary information.

The data analyzed in this study was obtained from the epidemiological investigation reports that were released by the Israeli Ministry of Health (Israeli Ministry of Health, 2020). From the total of 883 PCR laboratory confirmed COVID-19 cases we analyzed the 384 cases that were investigated epidemiologically, allowing us to separate the incoming from abroad (quarantined on arrival by policy) to local cases. This allowed us to separate the imported cases (travelers arriving from abroad) from the locally infected cases. The data spans over the first month of the COVID-19 outbreak in Israel, beginning in February 21^st^ 2020 and going until March 20^th^. We further note that during the examined period the number of PCR tests performed rose daily, reaching 1869 at the end of the period. However, the proportion of positive tests remained approximately the same, as 7.9% (Israeli Ministry of Health, 2020).

The incidence curve was modelled as a fit to an exponential growth function (de Silva, et al., 2009; Zhao, et al., 2020). Several serial interval distributions that were estimated for COVID-19 were examined (Nishiura, et al., 2020; Ganyani, et al., 2020; Zhao, et al., 2020).

## 3. Results

The dynamics of epidemic spread in Israel in terms of daily reported confirmed cases is shown in fig. 1. There are two distinct regimes in the curve. The first ended in March 12, 2020 in which the local to travel associated infection ratio was around 1:3 (up to 1:2.5), i.e., for every three imported cases, there was one local infection. The second period, began a day later. The first distinct sign for the change was evident on March 14^th^ were the ratio rose to 1:2, then continued to rise on March 17^th^ to 1:1.5 and finally, after a month into the epidemic in Israel, on March 21^st^ there were more local infections in Israel than imported cases (the ratio was above 1:1).

**Fig. 1.**
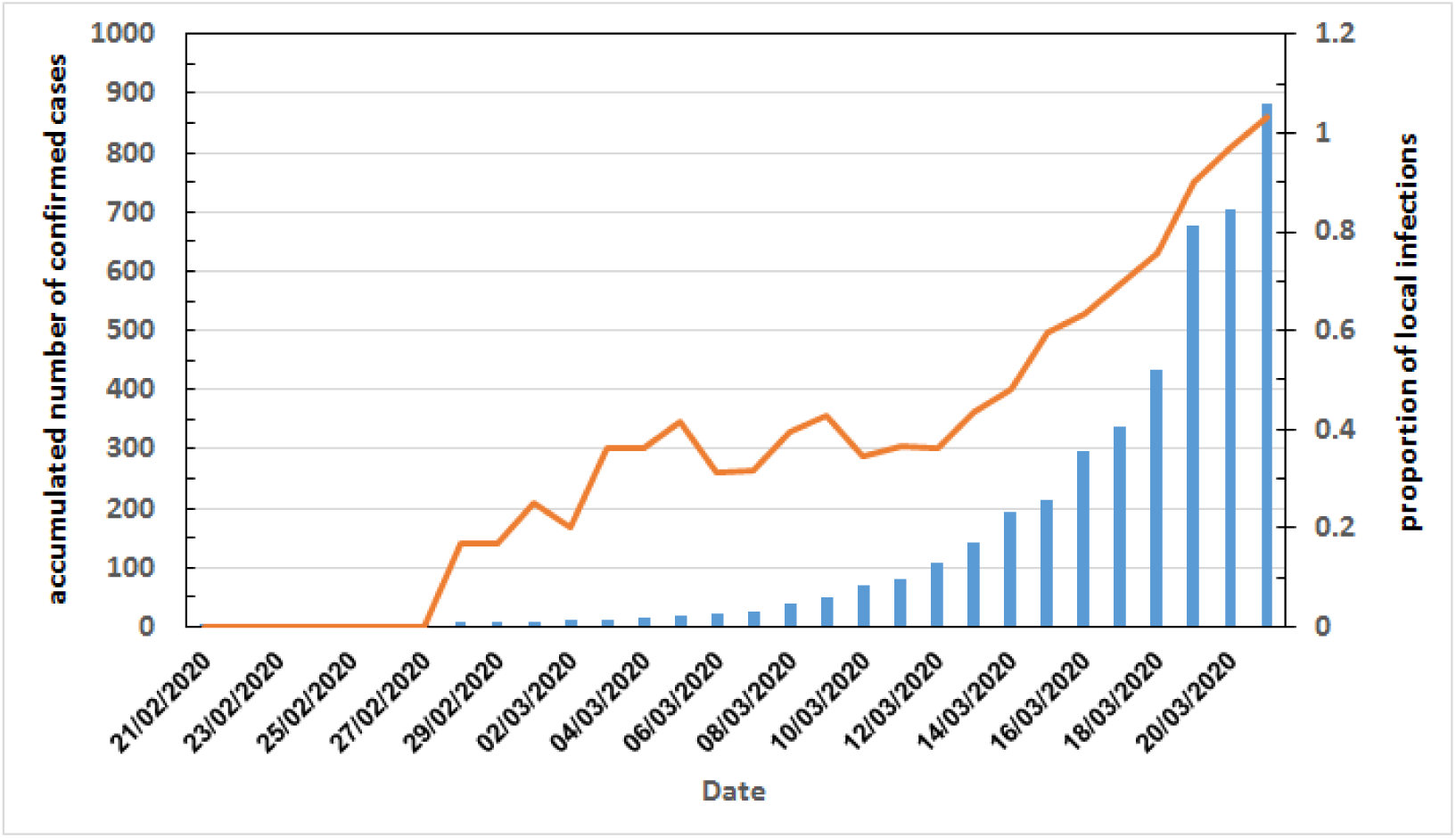
Accumulated reported confirmed COVID-19 cases in Israel (blue bars) and the ratio of local infections to travel associated infections (orange line)

March 12^th^ is two days after the Jewish holiday of Purim, which this year was celebrated on March 10^th^. Purim customs include wearing masks and costumes and holding public celebrations, parades as well as religious gatherings. Because Purim is also a school holiday, many celebration are held a day or two earlier.

The effective reproduction number, *R*_0_, was estimated for the daily new cases data of the first month of the COVID-19 epidemic. The lowest estimated *R*_0_ was 2.08 (95%CI 1.93–2.6) for the Gamma distributed serial interval with mean 4.4 and SD 3 days (Zhao, et al., 2020). The highest estimate was 2.37 (95%CI 2.16–2.61) for the Gamma distributed serial interval with mean 5.2 and SD 2.8 days (Ganyani, et al., 2020). The mean *R*_0_ overall the serial interval distributions examined, was 2.19.

The SEQIJR model solutions are characterized by 3 locally stable equilibrium points in parameters space. These refer to the following regimes or types of dynamics of epidemic spread: controlled (decaying), flattened, uncontrolled (baseline SEIR model; further details can be found in the supplementary material). In this study we simulated three scenarios, the first corresponding to the first equilibrium and two corresponding to the second:

1. A controlled high-efficiency quarantining (decaying green curve in Fig. 2). This regime is characterized by an early entry of asymptomatic suspected population to home quarantine. Moreover we assume the infectiousness in home quarantine is one sixth compared to free asymptomatic. The efficiency of isolation is 70%.
2. A flattened for medium-efficiency quarantining (purple curve in Fig. 2). This regime is characterized by a late entry of asymptomatic suspected population to home quarantine. Moreover we assume the infectiousness in home quarantine is one third compared to free asymptomatic. The efficiency of isolation is 70%.
3. A flattened for low-efficiency quarantining (light-blue curve in Fig. 2). This regime is characterized by a late entry of asymptomatic suspected population to home quarantine. Moreover we assume the infectiousness in home quarantine is similar to free asymptomatic. The efficiency of isolation is 30%.

As discussed above, the dynamics of epidemic spread in Israel until March 8, 2020 corresponds to the controlled regime characterizes with *R*_0_ of the order of half (green curve in Fig. 2). On the other hand, after March 8, 2020 the regime corresponds to the flattened regime with low-efficiency quarantining and *R*_0_ = 2.19.

**Fig. 2.**
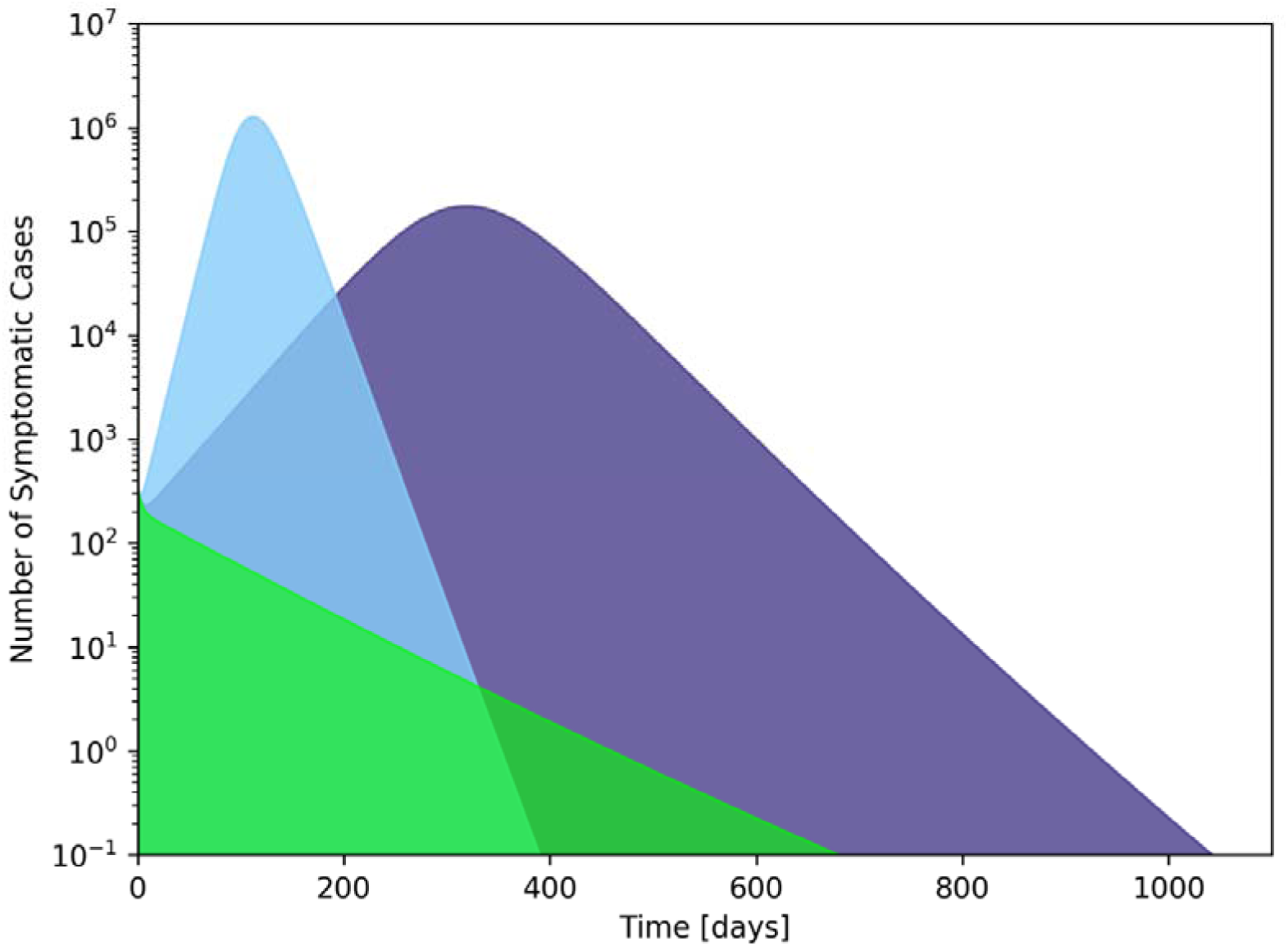
Total number of symptomatic cases in Israel from SEQIJR model simulations for 3 scenarios: green – high-efficiency quarantining, purple – medium-efficiency quarantining, light blue – low-efficiency quarantining.

## 4. Discussion

Israel has begun early its mitigation policy against the COVID-19 epidemic. From the beginning of February, it was decided: to close all border passages (via land, sea and air) to people that are not Israeli citizens or resident that have visited China recently; to stop all direct flights from Israel to China and to require Israeli citizens returning from China or that were in touch with a confirmed COVID-19 patients to a 14 days home quarantine. A short while later, the quarantine requirement was expanded to other Asian countries. The first COVID-19 patients in Israel were two passengers that returned from the “Diamond Princess”, in 21 and 23 of February. They entered directly to hospital isolation. A week later, a passenger from Italy was diagnosed as a COVID-19 patient, and Italy was added to the list of countries that require 14 days quarantine.

The quarantine-isolation policy succeeded in keeping the rate of daily new cases small, up until March 9^th^. Then, 4 days later, a sudden change of regime has occurred, which was manifested by the distinct change of the epidemic curve of Israel towards exponential growth. March 15^th^ marks the first time that the daily new cases of locally infected were higher than the new travel associated cases.

The timing of this abrupt change is not of coincidence. Regarding the cases arriving from abroad, a requirement for home quarantine affecting all travelers arriving began on Match 9^th^. Moreover, during the period between March 9^th^ to 11^th^ a Jewish holiday, Purim, was celebrated. This holiday is characterized by big parades organized by local municipalities, as well as religious gatherings and privately organized parties. Although authorities cancelled the public parades, many privately organized and religious crowding had occurred. Regrettably, these drove Israel from a controlled, mitigated regime to an exponential growth, as described in the results section. Therefore, despite its intense efforts, Israel’s effective *R*_0_ for the period ending in March 20^th^ stands on around 2.19, slightly smaller than the *R*_0_ of 2.6–3.2 estimated for the republic of Korea and Italy, for the period ending in March 5^th^. Such abrupt transition based on social behavior emphasizes the fragility of mitigation policies.

We therefore emphasis the importance of early fine-tuned but intense directives for social distancing and isolation measures. This study clearly demonstrates the lesson learned from the Israeli policy, that even a short lapse in public responsiveness can have a dramatic effect on public health during pandemic outbreak.

## Data Availability

Israel Ministry of Health. Information on confirmed patients and COVID-19 press releases. Accessed Mar 22, 2020.

https://govextra.gov.il/ministry-of-health/corona/corona-virus/spokesman-messages-corona/

